# Composite interventions on outcomes of severely and critically ill patients with COVID-19 in Shanghai, China

**DOI:** 10.1101/2023.05.10.23289325

**Authors:** Jiasheng Shao, Rong Fan, Chengnan Guo, Xuyuan Huang, Runsheng Guo, Fengdi Zhang, Jianrong Hu, Gang Huang, Liou Cao

## Abstract

**Background:** The sixty-day effects of initial composite interventions for the treatment of severely and critically ill patients with COVID-19 are not fully assessed.

**Methods:** Using a bayesian piecewise exponential model, we analyzed the 60-day mortality, health-related quality of life (HRQoL) and disability in 1082 severely and critically patients with COVID-19 between December 8, 2022 and February 9, 2023 in Shanghai, China. The final 60-day follow-up was completed on April 10, 2023.

**Results:** Among 1082 patients (mean age, 78.0 years), 421 [38.9%] women), 139 patients (12.9%) died within 60 days. Azvudine had a 99.8% probability of improving 2-month survival (adjusted HR, 0.44 [95% credible interval, 0.24-0.79]) and Paxlovid had a 91.9% probability of improving 2-month survival (adjusted HR, 0.71 [95% credible interval, 0.44-1.14]) compared with the control. IL-6 receptor antagonist, Baricitinib, and a-thymosin each had a high probability of benefit (99.5%, 99.4%, and 97.5%, respectively) compared to their controls, while the probability of trail-defined statistical futility (HR >0.83) was high for therapeutic anticoagulation (99.8%; HR, 1.64 [95% CrI, 1.06-2.50]), and glucocorticoid (91.4%; HR, 1.20 [95% CrI, 0.71-2.16]). Paxlovid, Azvudine and therapeutic anticoagulation showed significant reduction in disability (p<0.05)

**Conclusions:** Among severely and critically ill patients with COVID-19 who received 1 or more therapeutic interventions, treatment with Azvudine had a high probability of improved 60-day mortality compared with the control, indicating its potential in resource-limited scenario. Treatment with IL-6 receptor antagonist, Baricitinib, and a-thymosin also had high probabilities of benefit of improving 2-month survival, among which a-thymosin could improve HRQoL. Treatment with Paxlovid, Azvudine and therapeutic anticoagulation could significantly reduce disability at day 60.

## Introduction

In the end of 2022, China was inclined to change course and adopt a “living with COVID-19” strategy(1). Since then, COVID-19 infections have spread rapidly in major cities in China, including Shanghai, where the predominant SARS-CoV-2 variant, Omicron BF.7 has brought great pressure on the healthcare facilities. A model foresees that China’s outbreak will reach a first peak on 13 January with 3.7 million new cases per day and COVID-19-related deaths are expected to peak 10 days later at around 25,000 per day(2).

To date, majority of studies are mainly focused on the clinical characteristics and shorter-term progression of the viral infection in mild to moderate cases(3, 4). Huang et al. have described the one and two-year evolution of health outcomes in COVID-19 survivors, regardless of initial disease severity(5, 6). However, few studies with large sample sizes have specifically reported the clinical outcomes in severely and critically ill survivors with COVID-19. For severe and critical cases, cytokine storm is believed to be one of the major reasons of acute respiratory distress syndrome (ARDS) and multiple-organ failure which means comprehensive interventions should be administered in the early stage of the infection to reduce mortality(7). Some cohort research indicated critically ill patients who received antiviral agents, immune modulators, immunoglobulin, anticoagulation, antiplatelet and corticosteroids therapy have broad variations on the clinical outcome(8–13). For instance, antiviral agents such as nirmatrelvir-ritonavir (Paxlovid) and Molnupiravir that are currently available for the treatment of COVID-19 infection. These medications have been shown to reduce the risk of mortality in the post-acute phase in hospitalized patients(14).

However, limited data is available regarding the initial comprehensive interventions translating into clinical effects on survival, disability, and health-related quality of life (HRQoL), particularly for severely and critically ill patients with COVID-19(5). The aim of the study is to evaluate the effectiveness of these composite treatments on 60-day outcomes, including mortality, disability, and health-related quality of life (HRQoL) who receive one or more of these treatments.

## Methods

### Study Designs and Participants

During the COVID-19 pandemic, we conducted a retrospective, single-centered study involving 1082 severely and critically patients with COVID-19 which were confirmed by reverse transcriptase polymerase chain reaction (RT-PCR) or COVID-19 antigen test between December 8, 2022 and February 9, 2023 in Shanghai, China. The severe and critical illness of COVID-19 infection was defined by Guidelines on the Diagnosis and Treatment of COVID-19 (10th Trial Edition)(15). The final 60-day follow-up was completed on April 10, 2023.

The study was approved by the Ethics Committee of Jiading District Central Hospital Affiliated Shanghai University of Medicine & Health Sciences and performed (Approval code: 2023K15). Patients or their lawful caretakers provided written informed consent. The demographic profile (age, sex, pre-existing disorders) for enrolled participants, clinical course (diagnosis date, admission date, symptoms, the treatment recipes, disease severity) and vaccination doses information were extracted from medical records.

We divided all patients who received one or more interventions into six therapeutic domains: antivirals, immune modulators, intravenous immunoglobulin, antiplatelet, anticoagulation, and glucocorticoids, which were based a previous study and our national treatment guideline(9, 15). Briefly, patients in the antiviral domain received nirmatrelvir-ritonavir (Paxlovid), Azvudine (a nucleoside analog), or no antiviral medicines; patients in the immune modulation domain received tocilizumab (an IL-6 receptor antagonist), Baricitinib (a Janus kinase-JAK inhibitor), α-thymosin (a non-specific T cell activator) or no immune modulator; patients in the Intravenous immunoglobulin domain receives immunoglobulin for 3-5 days (given if clinical deterioration occurs); patients in the antiplatelet domain received aspirin, a P2Y12 inhibitor (clopidogrel or ticagrelor), or no antiplatelet therapy; patients in the anticoagulation domain received thrombo-prophylactic or therapeutic-dose anticoagulation with low molecular weight heparin (LMWH) or in accordance with usual administration; patients in the glucocorticoid domain received a (7–10)-day course of intravenous dexamethasone or methylprednisolone, or no glucocorticoids(15). The medications used in each domain were as follows: Paxlovid (nirmatrelvir 300 mg and ritonavir 100 mg twice per day for 5 consecutive days), Azvudine (5mg once per day for up to 14 days), tocilizumab (8mg/kg per day for 2 doses), Baricitinib (4mg per day for up to 14 day), α-thymosin (1.6 mg once per day for at least 7 consecutive days), intravenous immunoglobulin (20g per day for up to 5 days), antiplatelet (aspirin 100 mg once daily, clopidogrel 75 mg once daily, or ticagrelor 60 mg twice daily), anticoagulation (thrombo-prophylactic LMWH 4000 IU per day or therapeutic dose 100 IU/kg twice per day) and fixed-dose glucocorticoid (dexamethasone, 5mg per day or methylprednisolone 40 mg per day)(12, 15, 16). The flowchart of our cohort study was shown in **Figure 1**. The baseline characteristics of patients in one or more domains enrolled were presented in **Table 1.** Domain-specific inclusion and exclusion criteria for patients within each domain are shown in **Supplementary Table 1.**

**Figure 1.**
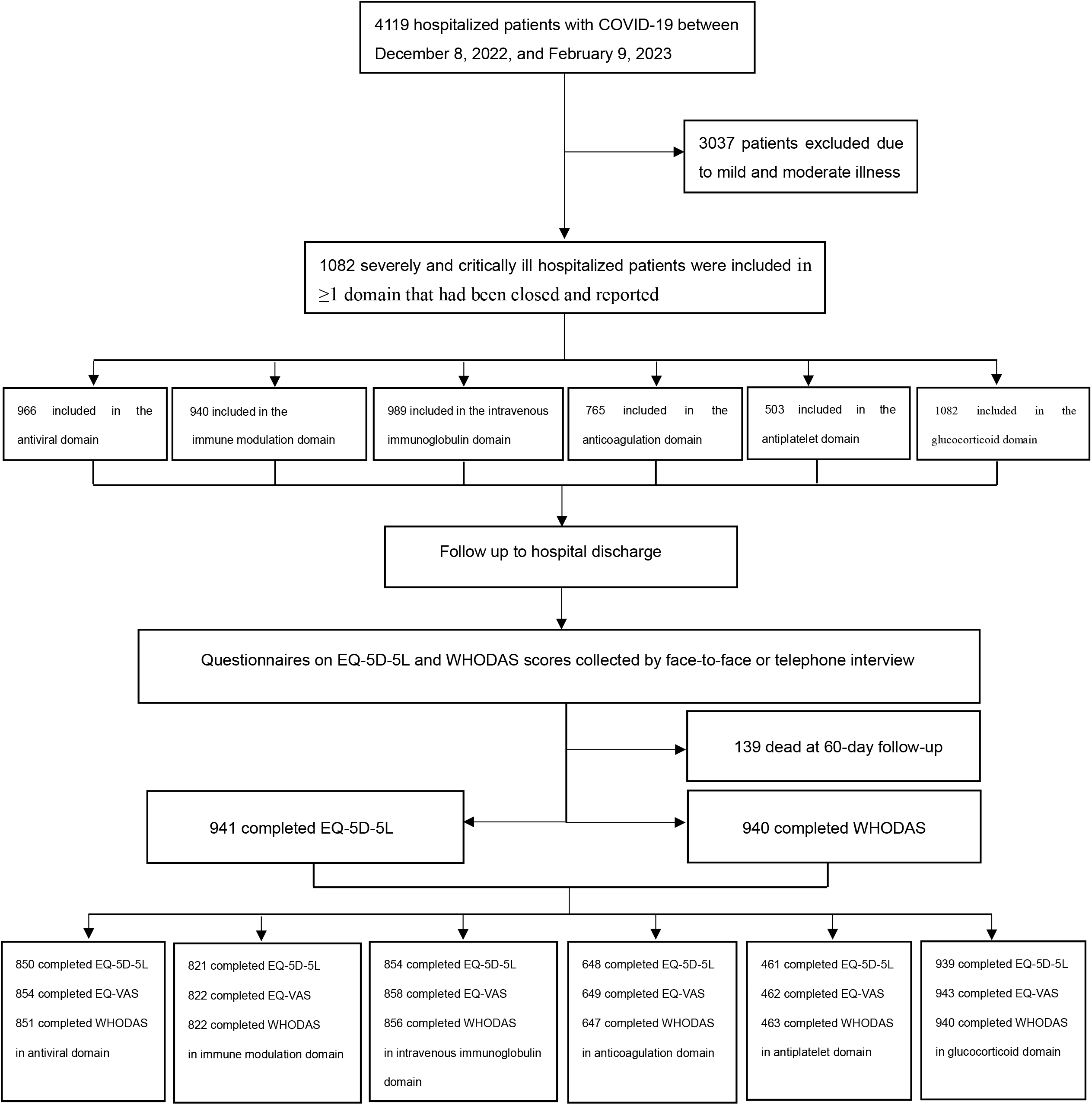
Flow chart of the study participants

**Table 1.**
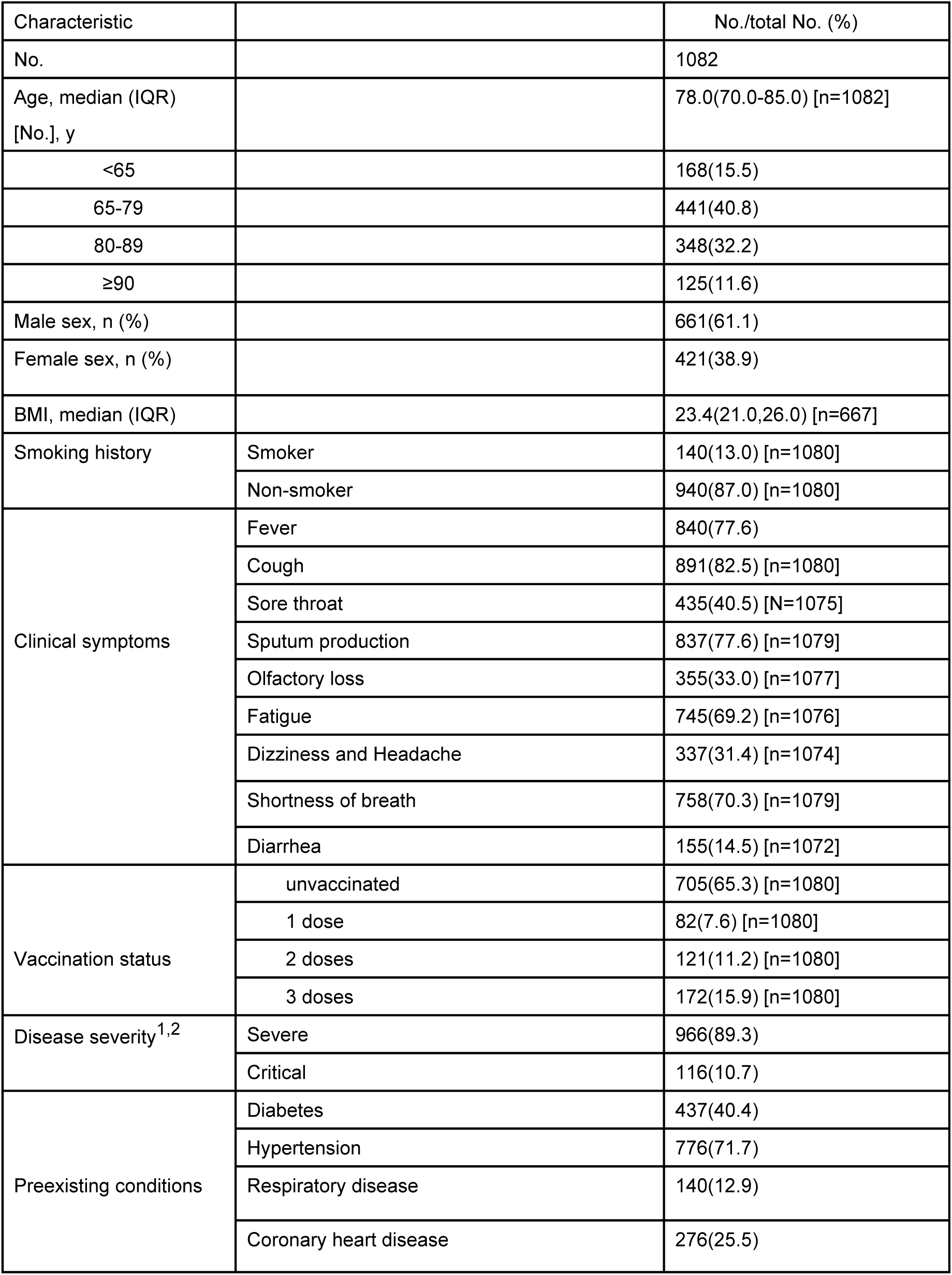
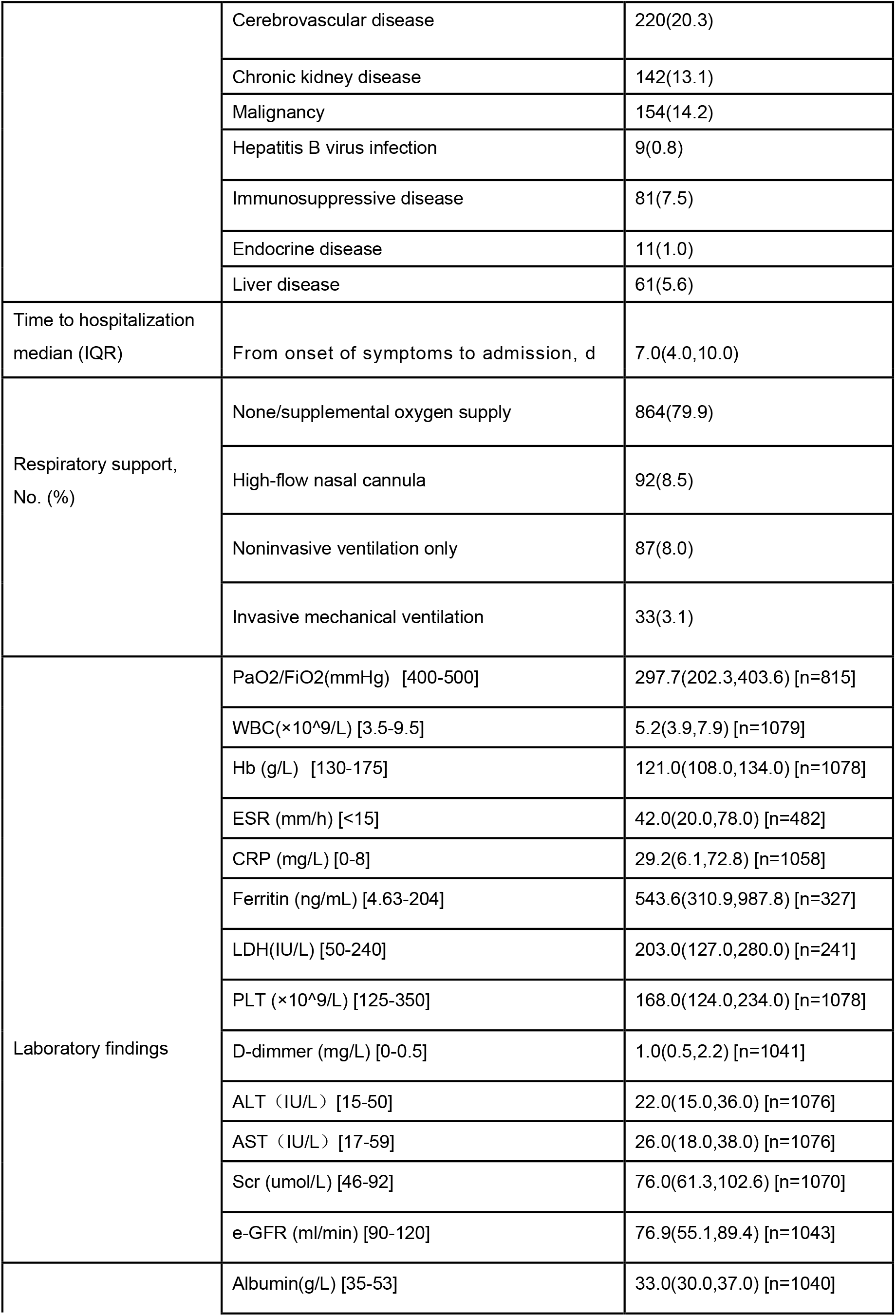
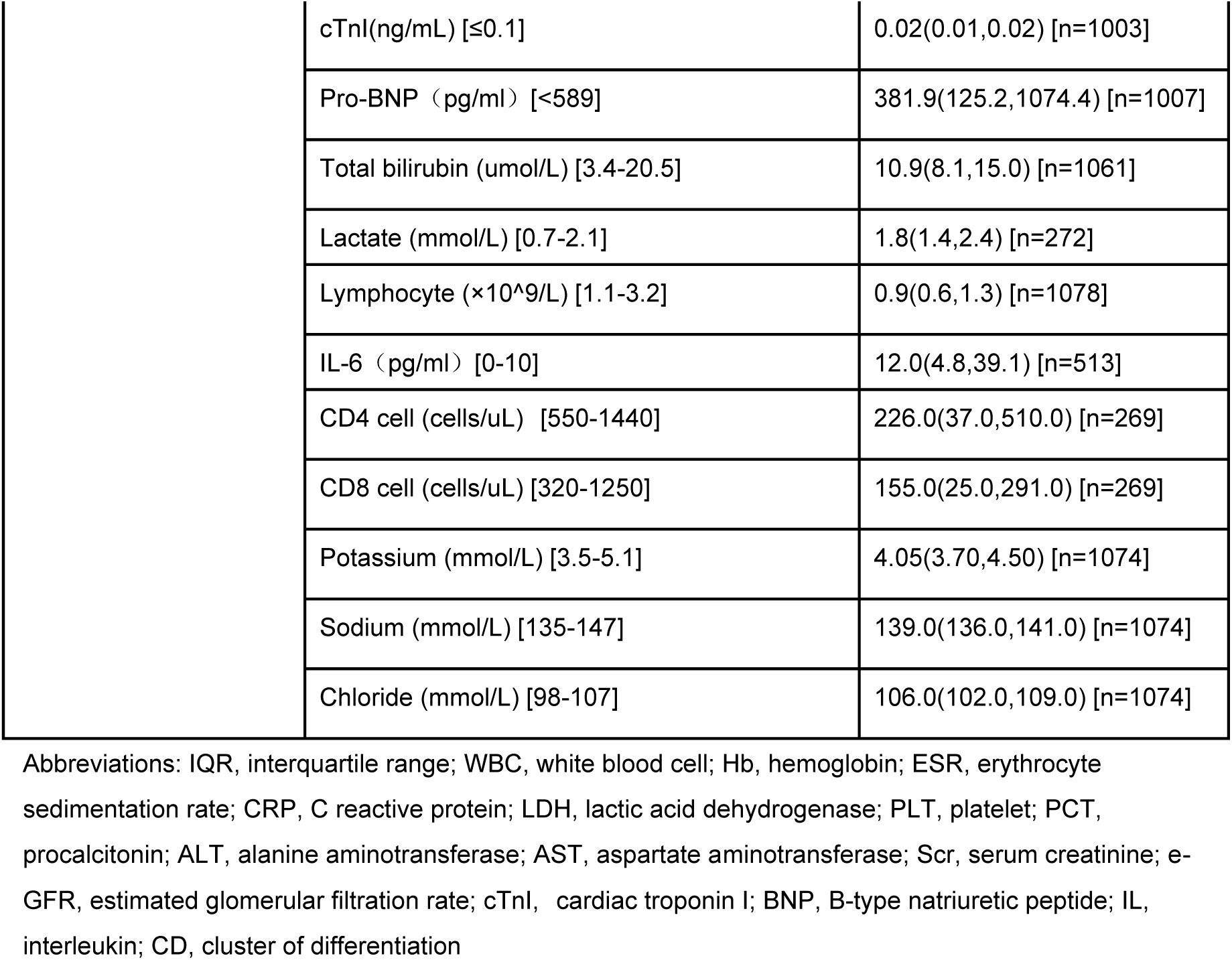
Baseline characteristics of patients in 1 or more domains enrolled.

| Characteristic |  | No./total No. (%) |
| --- | --- | --- |
| No. |  | 1082 |
| Age, median (IQR)<br>[No.], y |  | 78.0(70.0-85.0) [n=1082] |
| <65 |  | 168(15.5) |
| 65-79 |  | 441(40.8) |
| 80-89 |  | 348(32.2) |
| ≥90 |  | 125(11.6) |
| Male sex, n (%) |  | 661(61.1) |
| Female sex, n (%) |  | 421(38.9) |
| BMI, median (IQR) |  | 23.4(21.0,26.0) [n=667] |
| Smoking history | Smoker | 140(13.0) [n=1080] |
|  | Non-smoker | 940(87.0) [n=1080] |
| Clinical symptoms | Fever | 840(77.6) |
|  | Cough | 891(82.5) [n=1080] |
|  | Sore throat | 435(40.5) [N=1075] |
|  | Sputum production | 837(77.6) [n=1079] |
|  | Olfactory loss | 355(33.0) [n=1077] |
|  | Fatigue | 745(69.2) [n=1076] |
|  | Dizziness and Headache | 337(31.4) [n=1074] |
|  | Shortness of breath | 758(70.3) [n=1079] |
|  | Diarrhea | 155(14.5) [n=1072] |
| Vaccination status | unvaccinated | 705(65.3) [n=1080] |
|  | 1 dose | 82(7.6) [n=1080] |
|  | 2 doses | 121(11.2) [n=1080] |
|  | 3 doses | 172(15.9) [n=1080] |
| Disease severity<sup>1,2</sup> | Severe | 966(89.3) |
|  | Critical | 116(10.7) |
| Preexisting conditions | Diabetes | 437(40.4) |
|  | Hypertension | 776(71.7) |
|  | Respiratory disease | 140(12.9) |
|  | Coronary heart disease | 276(25.5) |
|  | Cerebrovascular disease | 220(20.3) |
|  | Chronic kidney disease | 142(13.1) |
|  | Malignancy | 154(14.2) |
|  | Hepatitis B virus infection | 9(0.8) |
|  | Immunosuppressive disease | 81(7.5) |
|  | Endocrine disease | 11(1.0) |
|  | Liver disease | 61(5.6) |
| Time to hospitalization median (IQR) | From onset of symptoms to admission, d | 7.0(4.0,10.0) |
| Respiratory support, No. (%) | None/supplemental oxygen supply | 864(79.9) |
|  | High-flow nasal cannula | 92(8.5) |
|  | Noninvasive ventilation only | 87(8.0) |
|  | Invasive mechanical ventilation | 33(3.1) |
| Laboratory findings | PaO<sub>2</sub>/FiO<sub>2</sub>(mmHg) [400-500] | 297.7(202.3,403.6) [n=815] |
| | WBC($\times 10^9$/L) [3.5-9.5] | 5.2(3.9,7.9) [n=1079] |
|  | Hb (g/L) [130-175] | 121.0(108.0,134.0) [n=1078] |
|  | ESR (mm/h) [<15] | 42.0(20.0,78.0) [n=482] |
|  | CRP (mg/L) [0-8] | 29.2(6.1,72.8) [n=1058] |
|  | Ferritin (ng/mL) [4.63-204] | 543.6(310.9,987.8) [n=327] |
|  | LDH(IU/L) [50-240] | 203.0(127.0,280.0) [n=241] |
| | PLT ($\times 10^9$/L) [125-350] | 168.0(124.0,234.0) [n=1078] |
|  | D-dimmer (mg/L) [0-0.5] | 1.0(0.5,2.2) [n=1041] |
|  | ALT (IU/L) [15-50] | 22.0(15.0,36.0) [n=1076] |
|  | AST (IU/L) [17-59] | 26.0(18.0,38.0) [n=1076] |
|  | Scr (umol/L) [46-92] | 76.0(61.3,102.6) [n=1070] |
|  | e-GFR (ml/min) [90-120] | 76.9(55.1,89.4) [n=1043] |
|  | Albumin(g/L) [35-53] | 33.0(30.0,37.0) [n=1040] |

|  |  |
| --- | --- |
| cTnI(ng/mL) [$\leq 0.1$] | 0.02(0.01,0.02) [n=1003] |
| Pro-BNP (pg/ml) [$< 589$] | 381.9(125.2,1074.4) [n=1007] |
| Total bilirubin (umol/L) [3.4-20.5] | 10.9(8.1,15.0) [n=1061] |
| Lactate (mmol/L) [0.7-2.1] | 1.8(1.4,2.4) [n=272] |
| Lymphocyte ($\times 10^9/L$) [1.1-3.2] | 0.9(0.6,1.3) [n=1078] |
| IL-6 (pg/ml) [0-10] | 12.0(4.8,39.1) [n=513] |
| CD4 cell (cells/uL) [550-1440] | 226.0(37.0,510.0) [n=269] |
| CD8 cell (cells/uL) [320-1250] | 155.0(25.0,291.0) [n=269] |
| Potassium (mmol/L) [3.5-5.1] | 4.05(3.70,4.50) [n=1074] |
| Sodium (mmol/L) [135-147] | 139.0(136.0,141.0) [n=1074] |
| Chloride (mmol/L) [98-107] | 106.0(102.0,109.0) [n=1074] |

Abbreviations: IQR, interquartile range; WBC, white blood cell; Hb, hemoglobin; ESR, erythrocyte sedimentation rate; CRP, C reactive protein; LDH, lactic acid dehydrogenase; PLT, platelet; PCT, procalcitonin; ALT, alanine aminotransferase; AST, aspartate aminotransferase; Scr, serum creatinine; e-GFR, estimated glomerular filtration rate; cTnI, cardiac troponin I; BNP, B-type natriuretic peptide; IL, interleukin; CD, cluster of differentiation

### Outcome Measures

The main outcome of our study was to assess all-cause mortality within 60 days. The secondary outcome included HRQoL at 60 days measured using the 5-level EuroQol-5 Dimension (EQ-5D-5L) utility score and visual analog scale (VAS) score, and disability level at 60 days measured using the 36-item World Health Organization Disability Assessment schedule (WHODAS) 2.0(17). Data was collected by face-to-face or telephone interview with the participants, their relatives, or health care professional in our hospital.

The EQ-5D-5L is a preference-based health-related quality of life (HRQoL) instrument comprised of five dimensions: mobility, self-care, usual activities, pain/discomfort, and anxiety/depression. Respondents are asked to choose the most appropriate option from five alternatives (no, slight, moderate, severe or extreme problems). Additionally, respondents were asked to indicate their present health state on a visual analogue scale (EQ VAS) ranging from the worst imaginable health state (“0”) to the best imaginable health state (“100”). EQ-5D-5L utility scores were calculated where a valid response (0 to 4) was available for each of the 6 EQ-5D-5L domains. Scores were calculated using the crosswalk link function and the individual responses to the EQ-5D-5L descriptive system, using the China time trade off (TTO) value set, with values between - 0.391 and 1.0(18).

The 36-item WHODAS 2.0 covers six domains of functioning with scores from 0 (no difficulty) to 4 (extreme difficulty) and a total score ranging from 0 to 144, with higher scores representing greater disability. The total score is divided by 144 and multiplied by 100 to convert it to a percentage of maximum disability. WHODAS percentage scores were used to determine mutually exclusive disability categories: 1) no disability (0–4.5%); 2) mild disability (4.5–24.5%); 3) moderate disability (24.5–49.5%); 4) severe disability (49.5–95.5%); and 5) complete disability (95.5–100%)(17).

### Statistical Analysis

The primary analysis was performed using a bayesian piecewise exponential model. The underlying hazard rate was piecewise constant for each 10-day period up to day 30 and the 30-day period from day 30 to day 60. The prior distribution for each hazard rate was a γ distribution with 1 day of exposure and a mean equal to the total exposure (in days) divided by the total number of events. The primary model estimated treatment effects (log hazard ratios [HRs]) for each intervention relative to control within each domain with standard normal priors. The primary model included variables for each domain with each domain treatment as a category (with control interventions from each domain set as the referent) and was adjusted for patient age (categorized into 4 groups), sex, smoking, disease severity, preexisting conditions, respiratory supports, and other treatments from other domains. The posterior distributions of the interventions’ HRs were summarized with medians, 95% credible intervals (CrIs), and the probability that an intervention was superior to the control for that domain (i.e., HR <1.0). Harm was defined as the probability the HR was greater than 1. Futility was defined as the probability that there was not more than a 20% relative improvement in outcome (HR >0.83). A prespecified interaction was modeled between antiplatelet therapy (pooled P2Y12 inhibitor and aspirin group) in the antiplatelet domain and therapeutic-dose heparin in the anticoagulation domain. Statistical thresholds based on posterior probabilities for superiority and harm were used for the primary outcome to determine trial stopping rules, but were not used to guide interpretation of other findings; rather, effect sizes along with posterior probabilities are presented for all analyses.

Sixty-day mortality was analyzed with a bayesian logistic regression model. The EQ-5D-5L utility score was analyzed with a 2-part/mixture model including 2 components: a continuous distribution of EQ-5D-5L utility scores for patients who survived to day 60 and a point mass at 0 for patients who died before day 60. The posterior distributions of the mean difference between treatment and control for EQ-5D-5L utility scores were summarized with medians, 95% CrIs, and the probability that an intervention was superior to the control for that domain (i.e., a mean difference less than 0). Treatment effects were estimated for all patients, along with estimates for survivors only. The EQ VAS score was reported using descriptive statistics only and WHODAS disability category was calculated with ordinal mixture mode(17).

## Results

In the main outcome of our study, we found that 139 out of 1082 patients (12.9%) died within 60-day. **Figure 2** and **Figure 3** illustrate Azvudine had a higher probability of benefit compared to their control group (99.8%), while Paxlovid had the probability of benefit of 91.9%. IL-6 receptor antagonist, baricitinib, and a-thymosin each had a higher probability of benefit (99.5%, 99.4%, and 97.5%, respectively) compared to their control groups. On the other hand, the probability of benefit for intravenous immunoglobulin, therapeutic anticoagulation, antiplatelet, and glucocorticoid compared to their control groups was 52.5%, 1.5%, 90.3%, and 26.1%, respectively. However, the probability of trail-defined statistical futility was high for therapeutic anticoagulation (99.8%; HR, 1.64 [95% CrI, 1.06-2.50]), glucocorticoid (91.4%; HR, 1.20 [95% CrI, 0.71-2.16]). More detailed information regarding the parameters is provided in the **Figure 3**

**Figure 2.**
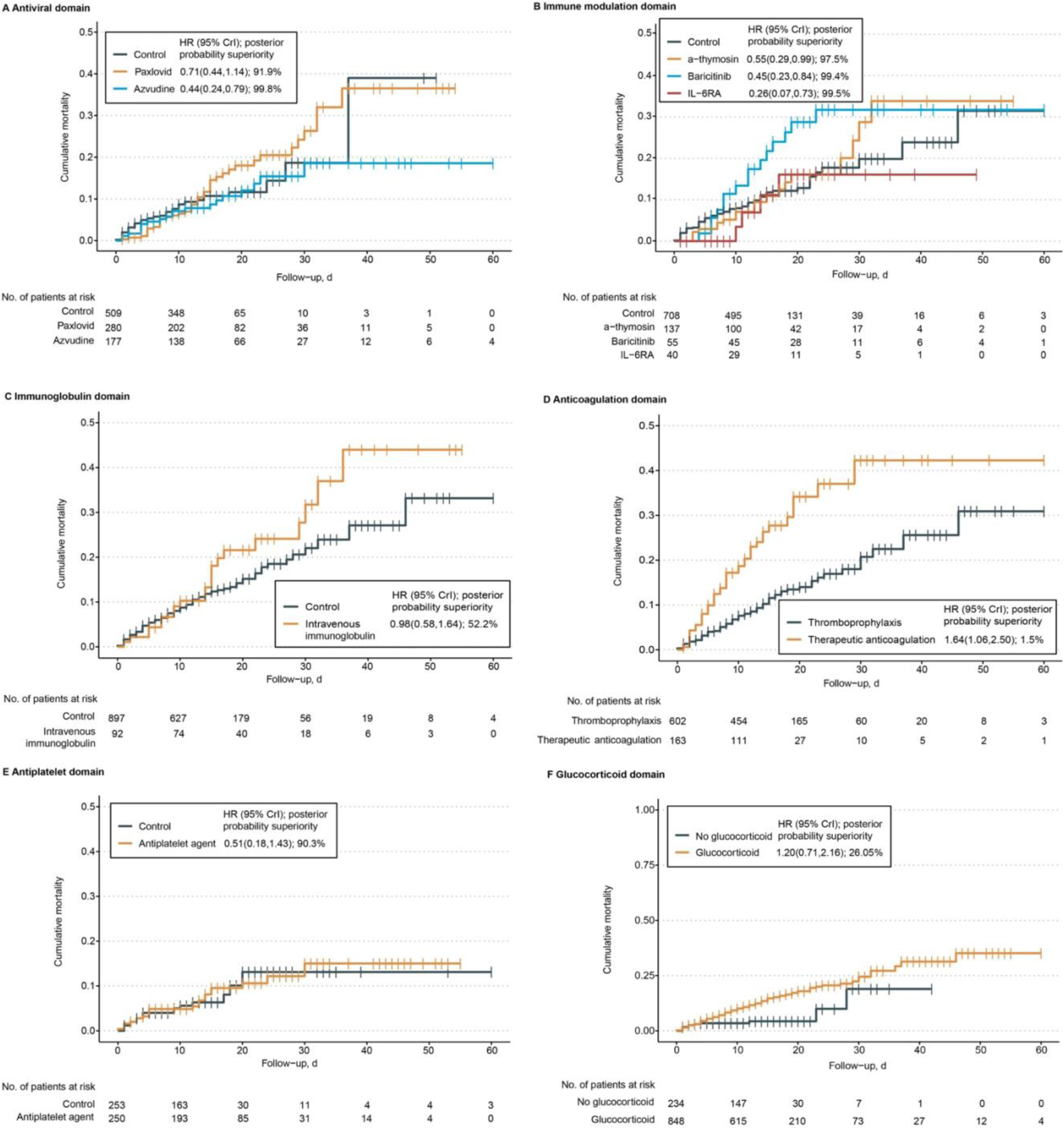
Kaplan-Meier curves for mortality through 60 days. The probability of superiority of each active intervention to control for 60-day mortality is reported from the fully adjusted bayesian model (adjusting for patient age, sex, smoking, disease severity, preexisting conditions, respiratory supports, and other treatments from other domains). Censored participants are indicated with vertical tick marks. CrI indicates credible interval.

**Figure 3.**
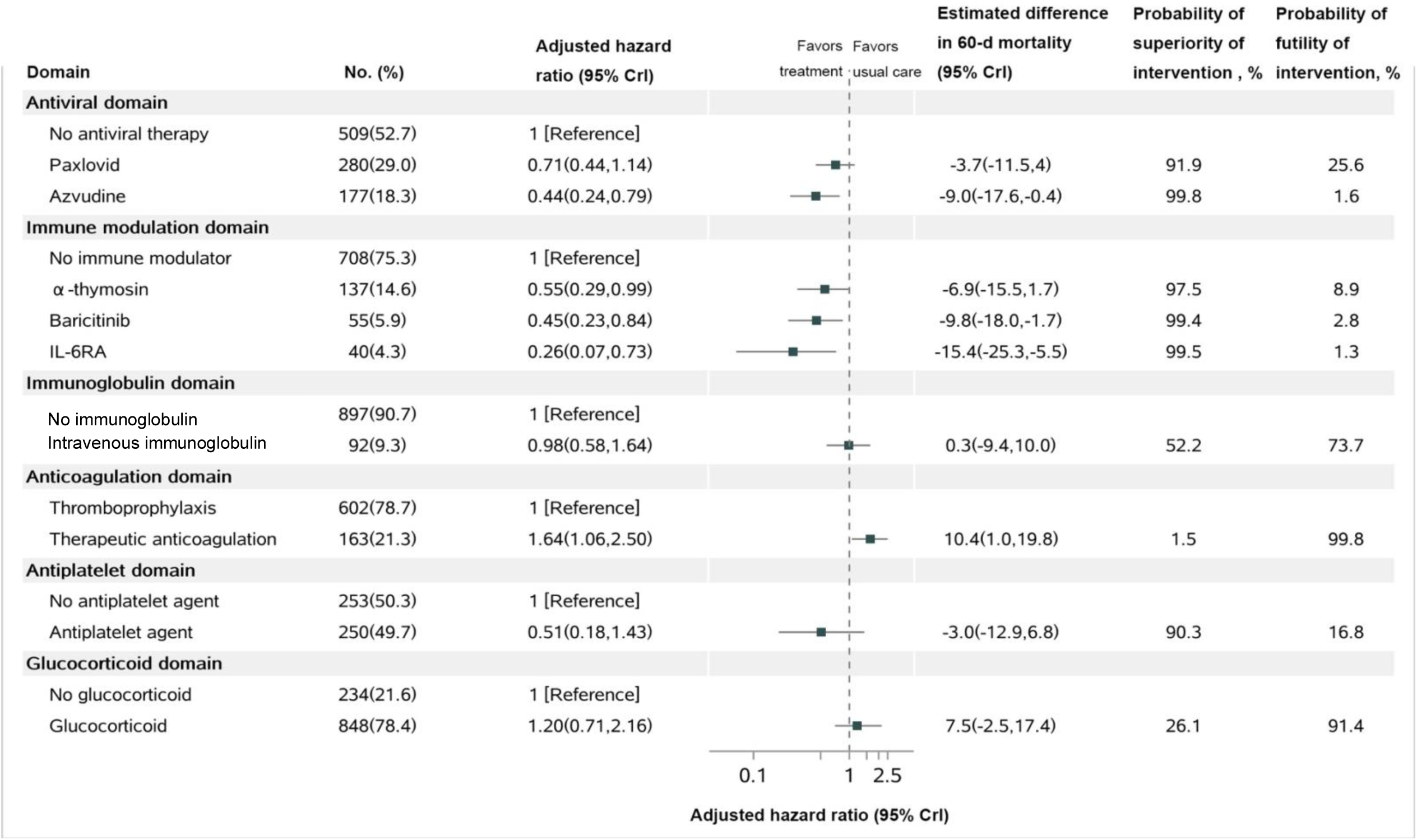
Mortality at 60 days. Notes: IL-6RA, interleukin-6 receptor antagonist; Hazard ratios <1 indicate improved survival and hazard ratios >1 indicate worsened survival. The difference in 60-day mortality is determined from the 60-day mortality rates which are estimated from the primary analysis model. For each domain, day 60 mortality rates are estimated for the population of patients divided within that domain based on their baseline covariates and the estimated model parameters. For each patient within the domain population, separate survival curves are predicted assuming the patient received each intervention within the domain. The mean of the survival curves was taken across patients to summarize the mean survival for each intervention within the domain population. The probability of superiority (hazard ratio <1) and futility (hazard ratio >0.83) is computed from a bayesian piecewise exponential model using the posterior distribution.

In the secondary outcomes, we assessed the EQ-5D-5L utility score, EQ VAS and WHODAS 2.0 score in 941 out of 943 survivors in follow-up (99.8%). In the overall observation, the median (IQR) EQ-5D-5L utility score in survivors was 0.64 (0.12-0.89) (n = 941) and the median (IQR) EQ VAS score was 70 (55–80) (n = 941). Mean EQ VAS scores in each domain are shown in **Supplementary Table 2**. Of the 940 survivors who completed WHODAS, 516 (54.89%) had moderate, severe, or complete disability at day 60. Notably, among these interventions, treatment with Paxlovid, Azvudine and therapeutic anticoagulation showed significant reduction in disability (p<0.05). Detailed information on disability in survivors is available in **Supplementary Table 3 and Table 4.**

The adjusted mean difference in the EQ-5D-5L utility score in the α-thymosin group was 0.08 (95% CrI, 0.01-0.16) units higher compared with the control, with a posterior probability of superiority of 97.7%; among all patients (survivors and non-survivors), the adjusted mean difference in the EQ-5D-5L utility score was also 0.08 (95% CrI, 0.00-0.16), with a probability of superiority of 97.6% (**Figure 4**). Based on the results, the mean EQ-5D-5L utility score in the Paxlovid group were lower than the control group, and the posterior probability of harm was 100.0% among survivors and all patients. Similarly, the mean EQ-5D-5L utility score in the Azvudine group was also lower compared to the control group, with a posterior probability of harm of 95.1% among survivors and 95.9% among all patients. The mean difference in EQ-5D-5L utility scores between each remaining intervention and their control group was presented in **Figure 4**.

**Figure 4.**
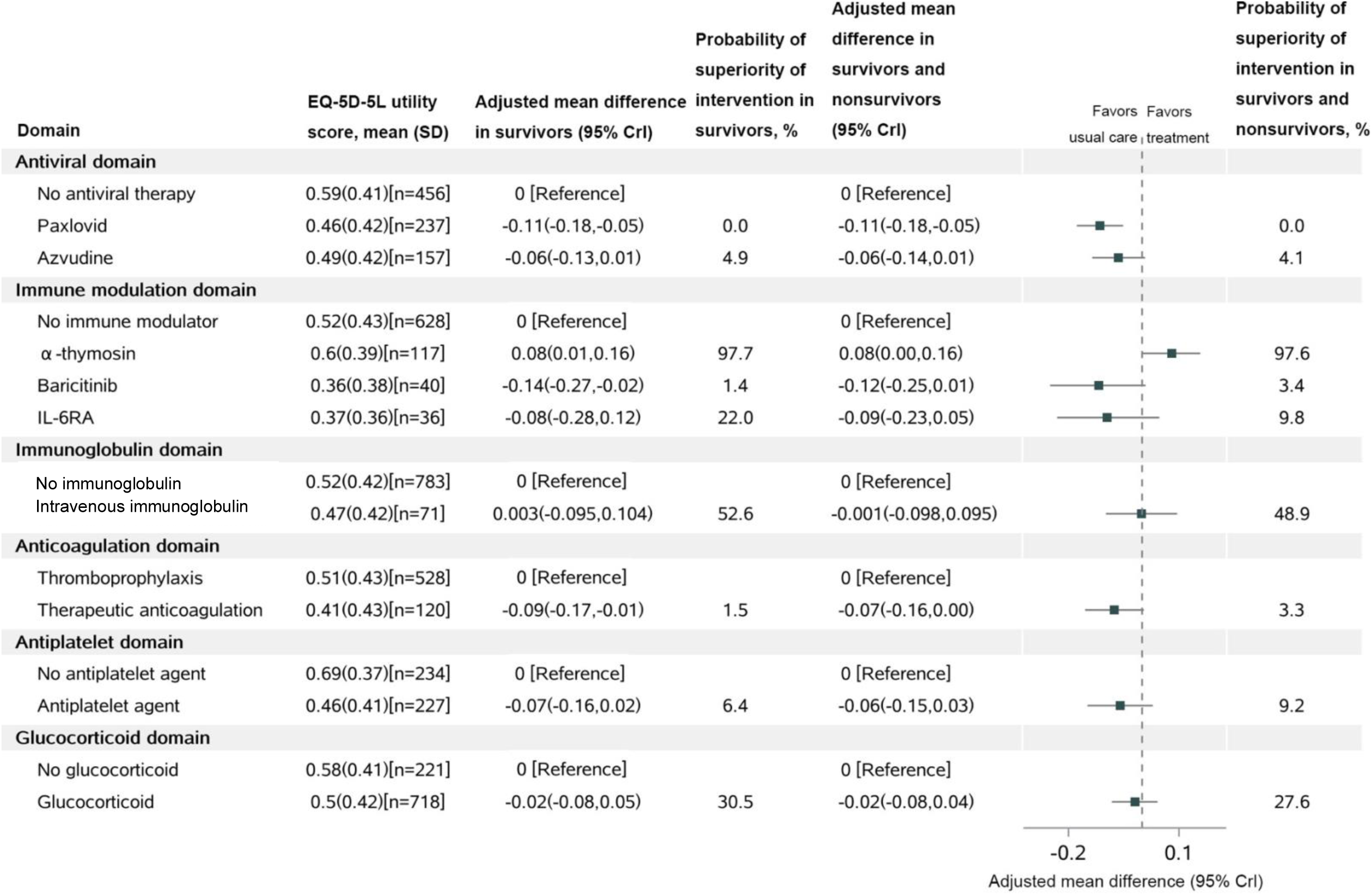
Health-related quality of life at 60 days. IL-6RA: IL-6 receptor antagonist; The probability of superiority and adjusted mean difference are computed from the posterior distribution of a bayesian 2-part/mixture model that multiply imputes 5-level EuroQol-5 Dimension (EQ-5D-5L) utility scores using patients’ baseline covariates. For patients who were known or imputed to be alive at 60 days, a value of EQ-5D-5L is multiply imputed from the continuous component of the 2-part/mixture model. For patients who were imputed as dead by 60 days, EQ-5D-5L was set to 0 and they did not contribute to the analysis of EQ-5D-5L in survivors.

## Discussion

In real-world practice, clinical interventions are administered in severely and critically ill patients aiming to increase long-term survival and improve HRQoL and functional status for survivors. However, most clinical studies in severely and critically ill patients have assessed shorter-term outcomes, which may not be patient-centered(19, 20). Longer-term trends after severe and critical illness such as hypoxemia, acute respiratory distress syndrome (ARDS) and septic shock, are characterized by frequent re-admission, persistent impairments in HRQoL and functional status, an elevated risk of mortality, and worsening of pre-exiting chronic disorders that may last for years following initial admission. Additionally, patients who initially survive may face late morbidity and mortality risks that perhaps outweigh the benefits of treatment. Therefore, the effectiveness of many regimens utilized in patients with severe and critical illness including COVID-19, remain unclear.

Paxlovid is recommended for mild to moderate COVID-19 cases within 5 days from symptoms onset(15). Liu et al. confirmed that it can be administered safely in severe adult patients with SARS-Cov-2 infection, but didn’t significantly reduce 28-day all-cause mortality and the duration of SARS–CoV-2 RNA clearance in these patients(21). Azvudine is a nucleoside analog that inhibits human immunodeficiency virus (HIV)-1 RNA-dependent RNA polymerase (RdRp), has also shown effectiveness in treating patients with COVID-19(22). Our study indicated Azvudine had a 99.8% of probability of improving survival over two months in COVID-19 patients with severe and critical illness when compared with the control. However, it did not improve HRQoL in survivors. Meanwhile, although Paxlovid showed a 91.9% superiority in terms of survival, the follow-up duration was not long enough to draw definitive conclusions.

Previous studies proved that IL-6 can activate Janus kinase-signal transducer and activator of transcription (JAK-STAT) pathway, induce inflammatory response and possibly form cytokine storm, which is an important factor for the development of ARDS and extrapulmonary organ damage, and IL-6 receptor antagonist can suppress the over-activation of human immune system(7). JAK inhibitors can reduce the inflammatory response induced by infection. A recent pilot study confirmed that treatment with Baricitinib plus standard of care (including use of corticosteroids) in critically ill patients with COVID-19 who were receiving invasive mechanical ventilation (IMV) or extracorporeal membrane oxygenation (ECMO) can reduce all-cause mortality at 28 days and 60 days(16). The two medicines are also recommended by our national guideline for the treatment of severely and critically ill patients with COVID-19(15, 23). In this observation cohort, both the IL-6 receptor antagonist and Baricitinib had higher probability of improving survival over 2 months.

Lymphocytopenia is a strong indicator of disease severity and prognosis in COVID-19 patients(24). Liu et al. reported that the treatment with α-thymosin can significantly reduce mortality in severe COVID-19 patients with severe lymphocytopenia by increasing T cell numbers and reversing T cell exhaustion(25). However, the therapeutic potential of α-thymosin remains controversial, with some studies reporting paradoxical results. In our investigation, α-thymosin, as a non-specific T cell activator, demonstrated a higher probability of improving both survival and HRQoL over 2 months. The observed discrepancies between studies could be attributed to differences in disease severity among patients(26).

Conversely, other treatment domains, including intravenous immunoglobulin and therapeutic anticoagulation, were found to be ineffective in improving patients’ 60-day mortality rate. It has been demonstrated that intravenous immunoglobulin has multifunctional immunomodulatory properties such as inhibiting the activation of the complement and proliferation of T helper 17 cell, neutralizing auto-antibodies, impairing the antigen presenting capabilities of dendritic cells, and expanding regulatory T cell populations. Therefore, it could be a good therapeutic strategy for hospitalized patients with COVID-19. A systematic review conducted by Liu et al. showed high-dose intravenous immunoglobulin (0.4-1.0 g/kg/day) may have a decreased risk for mortality in severe COVID-19 patients than patients with usual care(27). In contrast, Salehi et al. reported that the use of intravenous immunoglobulin did not reduce mortality in critically ill patients with COVID-19 without reducing mortality rate(28). This controversy in results could be attributed to differences in disease severity, dosage and timing of intravenous immunoglobulin treatment that affected its effects. Future work is need to identify the appropriate dosage, timing and subgroups of patients for treating severe and critical COVID-19 patients.

Previous clinical trials had also reported conflicting outcomes regarding the efficacy of therapeutic doses of LMWH in COVID-19 patients, with some studies manifesting beneficial effects and others showing no difference between therapeutic and prophylactic doses of LMWH. A randomized trial has indicated that therapeutic-dose anticoagulant may not provide any benefit to critically ill COVID-19 patients, which is consistent with our findings. The timing of anticoagulation initiation and the effect of anticoagulation may vary depending on the severity of the illness, which could be a possible explanation for the variability in outcomes reported previously(11).

Venous and arterial thromboembolism are commonly seen in severe/critically ill COVID-19 patients which is caused by platelet activation. Antiplatelet treatment can not only halt thrombosis, but also can alleviate the inflammatory response in these patients(29). However, the results of published studies on the effects of aspirin and P2Y12 antagonists (clopidogrel, ticagrelor) on the hospitalized patients with COVID-19 are still controversial(30). In our report, antiplatelet agents were found to have a 90.3% probability of improving 60-day survival rate, although there was no significant improvement in HRQoL among survivors at two months. The possible reason for this discrepancy could be whether there is a difference between patients who received treatment with antiplatelet before admission and those who were assigned to antiplatelet agents after admission. We need more randomized clinical trials with larger samples to explore whether pre-existing or other antiplatelet agents might be beneficial in COVID-19 infection.

There are clinical trials confirming that dexamethasone can lower in-hospital mortality among those who were receiving either IMV or oxygen alone(13, 19). However, in our study, glucocorticoid therapy showed a 26.1% probability of 60-day survival rate and a 30.5% probability of HRQoL improvement, indicating it did not improve survival or HRQoL. Of note, the patients who received glucocorticoid treatment were older than 70-year in our study, and this age difference may have contributed to the lack of efficacy for the regimen(23). Further research is needed to elucidate the efficacy of glucocorticoid therapy in COVID-19 patients of different age groups.

To the best of our knowledge, this is the first real-world observational research that has reported on the effect of composite treatments for COVID-19 on longer-term mortality, HRQoL, and disability in severely and critically ill patients after the ending of dynamic zero-COVID policy in Chinese population. This is noteworthy as recent studies have indicated that drugs such as Paxlovid and remdesivir did not show significant reduction in the risk of all-cause mortality in severely or critically ill adult patients with SARS-Cov-2 infection(8, 21). As the first domestic oral anti-COVID-19 drug launched in China, Azvudine has already been included in the national Guidelines on the Diagnosis and Treatment of COVID-19 (10th Trial Edition) for the treatment of adult patients with COVID-19 infections(31). Our results support the inclusion of Azvudine in the guideline for treating severely and critically ill COVID-19 patients, and highlights its potential in developing areas where access to Paxlovid is restricted.

There are some limitations to our study that should be addressed. Firstly, the duration of follow-up may not long enough to fully evaluate the long-term effectiveness of treatment in each domain. Secondly, the results may be influenced by the effects of interventions on different variant of COVID-19 and different vaccination status. Thirdly, we were unable to collect baseline HRQoL and disability scores during the severe or critical stage of patients upon admission, which limits our ability to assess changes in these scores over time. Fourthly, the number of patients who received IL-6 receptor antagonist in immune modulation domain was small, which may have introduced bias into the survival analysis. Finally, our study was conducted in a single center in China, and the results may not be generalizable to other settings or populations. However, we continue to follow up with these discharged patients and collect information on their survival rate and functional status to determine whether the observed effects are maintained over the long term.

## Data Availability

All data produced in the present study are available upon reasonable request to the authors

## Author contributions

Conceptualization, J.S., L.C. and G.H.; methodology, J.S., C.G. and R.F.; software, C.G., R.F. and X.H.; validation, R.G., X.H., Y.J. and H.L.; formal analysis, J.S., R.F., and C.G.; investigation, J.S., F.Z. and R.G.; resources, J.S., L.C. and G.H.; data curation, J.S.,C.G. and R.F.; writing—original draft preparation, J.S., C. G. and R.F.; writing-review and editing, J.S., R.F.; visualization, J.S. and C.G.; supervision, L.C. and G.H.; project administration, J.S., F.Z., R.G. and J. H.; funding acquisition, G.H. and L.C. All authors have read and agreed to the published version of the manuscript.

## Ethical approval Statement

This study was approved by the Ethics Committee of Jiading District Central Hospital affiliated to Shanghai University of Medicine and Health Sciences (Approval code 2023K15)

## Data Availability

All data produced in the present study are available upon reasonable request to the authors

## Acknowledgments

We acknowledge all patients involved in the study and their families. We thank Xiaofei Peng, Weilong Pan, Ben Yuan, Fenfang Yu, Caihe Wang for data collection from medical records. We would also like to thank Liping Wang, Hua Cheng, Chunying Han, Chuan Liu, Huaying Qiu for this face-to-face or telephone interview in our clinic. Finally, we thank Qiang Guo, Min Dai, Limin Tan, Manxiang Wang, Jiayan Liu, Guanghui Li for their medical advices and consultations.

## References

1. Ioannidis JPA, Zonta F, Levitt M. Estimates of COVID-19 deaths in Mainland China after abandoning zero COVID policy. Eur J Clin Invest. 2023:e13956.

2. Dyer O. Covid-19: China stops counting cases as models predict a million or more deaths. BMJ. 2023;380:2.

3. Wang B, Yu Y, Yu Y, Wang N, Chen F, Jiang B, et al. Clinical features and outcomes of hospitalized patients with COVID-19 during the Omicron wave in Shanghai, China. J Infect. 2023;86(1):e27–e9.

4. Shao J, Fan R, Hu J, Zhang T, Lee C, Huang X, et al. Clinical Progression and Outcome of Hospitalized Patients Infected with SARS-CoV-2 Omicron Variant in Shanghai, China. Vaccines (Basel). 2022;10(9).

5. Huang L, Yao Q, Gu X, Wang Q, Ren L, Wang Y, et al. 1-year outcomes in hospital survivors with COVID-19: a longitudinal cohort study. Lancet. 2021;398(10302):747–58.

6. Huang L, Li X, Gu X, Zhang H, Ren L, Guo L, et al. Health outcomes in people 2 years after surviving hospitalisation with COVID-19: a longitudinal cohort study. Lancet Respir Med. 2022;10(9):863–76.

7. Ye Q, Wang B, Mao J. The pathogenesis and treatment of the Cytokine Storm’ in COVID-19. J Infect. 2020;80(6):607–13.

8. Cilloniz C, Motos A, Castaneda T, Gabarrus A, Barbe F, Torres A, et al. Remdesivir and survival outcomes in critically ill patients with COVID-19: A multicentre observational cohort study. J Infect. 2023;86(3):256–308.

9. Writing Committee for the R-CAPI, Higgins AM, Berry LR, Lorenzi E, Murthy S, McQuilten Z, et al. Long-term (180-Day) Outcomes in Critically Ill Patients With COVID-19 in the REMAP-CAP Randomized Clinical Trial. JAMA. 2023;329(1):39–51.

10. Writing Committee for the R-CAPI, Estcourt LJ, Turgeon AF, McQuilten ZK, McVerry BJ, Al-Beidh F, et al. Effect of Convalescent Plasma on Organ Support-Free Days in Critically Ill Patients With COVID-19: A Randomized Clinical Trial. JAMA. 2021;326(17):1690–702.

11. Investigators R-C, Investigators AC-a, Investigators A, Goligher EC, Bradbury CA, McVerry BJ, et al. Therapeutic Anticoagulation with Heparin in Critically Ill Patients with Covid-19. N Engl J Med. 2021;385(9):777–89.

12. Investigators R-CWCftR-C, Bradbury CA, Lawler PR, Stanworth SJ, McVerry BJ, McQuilten Z, et al. Effect of Antiplatelet Therapy on Survival and Organ Support-Free Days in Critically Ill Patients With COVID-19: A Randomized Clinical Trial. JAMA. 2022;327(13):1247–59.

13. Group RC, Horby P, Lim WS, Emberson JR, Mafham M, Bell JL, et al. Dexamethasone in Hospitalized Patients with Covid-19. N Engl J Med. 2021;384(8):693–704.

14. Wan EYF, Wang B, Mathur S, Chan CIY, Yan VKC, Lai FTT, et al. Molnupiravir and nirmatrelvir-ritonavir reduce mortality risk during post-acute COVID-19 phase. J Infect. 2023.

15. National Health Commission of the People’s Republic of China. Diagnosis and treatment plan for COVID-19 (trial version 10). in Chinese. 2022.

16. Ely EW, Ramanan AV, Kartman CE, de Bono S, Liao R, Piruzeli MLB, et al. Efficacy and safety of baricitinib plus standard of care for the treatment of critically ill hospitalised adults with COVID-19 on invasive mechanical ventilation or extracorporeal membrane oxygenation: an exploratory, randomised, placebo-controlled trial. Lancet Respir Med. 2022;10(4):327–36.

17. TB Üstün NK, S Chatterji, J Rehm. Measuring Health and Disability, Manual for WHO Disability Assessment Schedule, WHODAS 2.0. 2010.

18. Luo N, Liu G, Li M, Guan H, Jin X, Rand-Hendriksen K. Estimating an EQ-5D-5L Value Set for China. Value Health. 2017;20(4):662–9.

19. Martinez-Guerra BA, Gonzalez-Lara MF, Roman-Montes CM, Tamez-Torres KM, Dardon-Fierro FE, Rajme-Lopez S, et al. Outcomes of patients with severe and critical COVID-19 treated with dexamethasone: a prospective cohort study. Emerg Microbes Infect. 2022;11(1):50–9.

20. Jimenez-Mora MA, Varela AR, Meneses-Echavez JF, Bidonde J, Angarita-Fonseca A, Siemieniuk RAC, et al. Patient-important outcomes reported in randomized controlled trials of pharmacologic treatments for COVID-19: a protocol of a META-epidemiological study. Syst Rev. 2021;10(1):289.

21. Liu J, Pan X, Zhang S, Li M, Ma K, Fan C, et al. Efficacy and safety of Paxlovid in severe adult patients with SARS-Cov-2 infection: a multicenter randomized controlled study. Lancet Reg Health West Pac. 2023:100694.

22. Zhang JL, Li YH, Wang LL, Liu HQ, Lu SY, Liu Y, et al. Azvudine is a thymus-homing anti-SARS-CoV-2 drug effective in treating COVID-19 patients. Signal Transduct Target Ther. 2021;6(1):414.

23. Chinese Thoracic S, Chinese Association of Chest Physicians Critical Care G. [Expert consensus on treatment of severe COVID-19 caused by Omicron variants]. Zhonghua Jie He He Hu Xi Za Zhi. 2023;46(2):101–10.

24. Xu B, Fan CY, Wang AL, Zou YL, Yu YH, He C, et al. Suppressed T cell-mediated immunity in patients with COVID-19: A clinical retrospective study in Wuhan, China. J Infect. 2020;81(1):e51–e60.

25. Liu Y, Pan Y, Hu Z, Wu M, Wang C, Feng Z, et al. Thymosin Alpha 1 Reduces the Mortality of Severe Coronavirus Disease 2019 by Restoration of Lymphocytopenia and Reversion of Exhausted T Cells. Clin Infect Dis. 2020;71(16):2150–7.

26. Bellet MM, Renga G, Pariano M, Stincardini C, D’Onofrio F, Goldstein AL, et al. COVID-19 and beyond: Reassessing the role of thymosin alpha1 in lung infections. Int Immunopharmacol. 2023;117:109949.

27. Liu X, Zhang Y, Lu L, Li X, Wu Y, Yang Y, et al. Benefits of high-dose intravenous immunoglobulin on mortality in patients with severe COVID-19: An updated systematic review and meta-analysis. Front Immunol. 2023;14:1116738.

28. Salehi M, Barkhori Mehni M, Akbarian M, Fattah Ghazi S, Khajavi Rad N, Moradi Moghaddam O, et al. The outcome of using intravenous immunoglobulin (IVIG) in critically ill COVID-19 patients’: a retrospective, multi-centric cohort study. Eur J Med Res. 2022;27(1):18.

29. Spaetgens B, Nagy M, Ten Cate H. Antiplatelet Therapy in Patients With COVID-19-More Is Less? JAMA. 2022;327(3):223–4.

30. Bolek T, Samos M, Jurica J, Stanciakova L, Pec MJ, Skornova I, et al. COVID-19 and the Response to Antiplatelet Therapy. J Clin Med. 2023;12(5).

31. Yu B, Chang J. The first Chinese oral anti-COVID-19 drug Azvudine launched. Innovation (Camb). 2022;3(6):100321.

