## Supplemental file for "Composite interventions on outcomes of severely and critically ill patients with COVID-19 in Shanghai, China"

### Supplementary material

**Table 1: Domain-specific Inclusion and Exclusion Criteria (1,2)**

| Domain | Inclusion criteria | Exclusion criteria |
|---|---|---|
| **Antiviral** | • Active COVID-19 infection with cycle threshold (Ct) value <30 times | • 30 ml/min≤creatinine clearance<60ml/min<br>• Severe liver dysfunction (Child-Pugh C)<br>• known or suspected pregnancy will result in exclusion from any intervention that include Paxlovid or Azvudine<br>• Known hypersensitivity to an agent specified as an intervention in this domain will exclude a patient from receiving that agent<br>• Known HIV infection will exclude a patient from receiving Paxlovid or Azvudine<br>• Known hypersensitivity to Paxlovid or Azvudine<br>•Receiving Salmeterol, Rifampicin, Tacrolimus, Sirolimus, Domperidone, Simvastatin, Rivaroxaban, Estazolam, Atorvastatin, Amiodarone, Propafenone, Carbamazepine as usual medications prior to this hospitalization or any administration of drugs mentioned above within 72 hours prior to assessment of eligibility will exclude a patient from receiving Paxlovid |

| | | |
|---|---|---|
| **Immune modulation** | • Active COVID-19 infection<br>• The level of IL-6 in serum was 100pg/ml and more (10 times more than upper limit of reference)<br>• C-reactive protein≥75 mg/L | • Patient has already received any dose of one or more of any form of α-thymosin, Baricitinib or Tocilizumab during this hospitalization or is on long-term therapy with any of these agents prior to this hospital admission<br>• Known condition or treatment resulting in ongoing immune suppression including neutropenia prior to this hospitalization<br>• The treating clinician believes that participation in the domain would not be in the best interests of the patient<br>• Known hypersensitivity to an agent specified as an intervention in this domain<br>• Known or suspected pregnancy will result in exclusion from α-thymosin, Tocilizumab and Baricitinib interventions.<br>• A baseline alanine aminotransferase or an aspartate aminotransferase that is more than five times the upper limit of normal will result in exclusion from receiving Tocilizumab<br>• A baseline platelet count < 50x10^9/L, or neutrophil < 0.5 x10^9/L will result in exclusion from receiving Tocilizumab<br>• Active tuberculosis, malignant tumor, thrombus, pregnancy will result in exclusion from receiving Baricitinib |

| | | |
|---|---|---|
| | | • Patients who suffered from thymoma or who received organ transplantation will result in exclusion from receiving Baricitinib |
| **Intravenous immunoglobulin** | • Active COVID-19 infection | • More than 14 days have elapsed since hospital admission<br>• Patient has already received treatment with any non-trial prescribed antibody therapy (monoclonal antibody, hyperimmune immunoglobulin, or convalescent plasma) intended to be active against COVID-19 during this hospital admission<br>•The treating clinician believes that participation in the domain would not be in the best interests of the patient<br>• Known hypersensitivity to an agent specified as an intervention in this domain<br>• Known previous history of transfusion-related acute lung injury<br>• Known objection to receiving plasma products |
| **Antiplatelet** | • Pre-exiting disorders need antiplatelet before COVID-19 infection.<br>• Recommended for all patients with severe and critical illness | • Clinical or laboratory bleeding risk or both that is sufficient to contraindicate antiplatelet therapy<br>• Patient is already receiving antiplatelet therapy or non-steroidal anti-inflammatory drug (NSAID) or a clinical decision has been made to commence antiplatelet or NSAID therapy<br>• Patients otherwise eligible for the therapeutic anticoagulation domain will be excluded from the |

| | | |
|---|---|---|
| | | antiplatelet domain if age is more than 75 years<br>• Creatinine clearance <30ml/min, or receiving renal replacement therapy or ECMO<br>• Known hypersensitivity to an agent specified as an intervention in this domain will exclude a patient from receiving that agent<br>•Known or suspected pregnancy will result in exclusion from the P2Y12 inhibitor intervention<br>•Administration or intention to administer Paxlovid will result in exclusion from the P2Y12 inhibitor intervention that are using ticagrelor as the P2Y12 inhibitor |
| **Anticoagulation** | • Pre-exiting disorders need anticoagulation therapy before COVID-19 infection<br>• Patients otherwise eligible for the therapeutic antiplatelet domain will be included if they already have received Paxlovid | • Clinical or laboratory bleeding risk or both that is sufficient to contraindicate therapeutic anticoagulation, including intention to continue or commence dual anti-platelet therapy<br>• Known or suspected previous adverse reaction to low molecular weight heparin (LMWH) including heparin-induced thrombocytopenia (HIT).<br>• Patients with a platelet count of less than 50 × 10^9/L, hemoglobin level below 80 g/L, a bleeding history within the past 30 days, or a creatinine clearance of less than 30 ml/min will be excluded in therapeutic anticoagulation group<br>• Patients with a platelet count of less than 50 × 10^9/L, a recent history of brain bleeding, or active |

| | | |
|---|---|---|
| | | bleeding who need more than 400ml of blood transfusion, or a creatinine clearance of less than 30 ml/min will be excluded in thromboprophylaxis group |
| **Glucocorticoid** | • PaO2/FiO2 and imaging of Chest CT deteriorated over time during hospitalization | •Known hypersensitivity to dexamethasone or methylprednisolone<br>• An indication to prescribe systemic glucocorticoid for a reason that is unrelated to the current episode, such as chronic corticosteroid use before admission, acute severe asthma, or suspected or proven *Pneumocystis jiroveci* pneumonia |

**Reference**


1. Chinese Thoracic S, Chinese Association of Chest Physicians Critical Care G. [Expert consensus on treatment of severe COVID-19 caused by Omicron variants]. Zhonghua Jie He He Hu Xi Za Zhi 2023;46:101-10.
2. National Health Commission of the People’s Republic of China. Diagnosis and treatment plan for COVID-19 (trial version 10). in Chinese. 2022.

**Table 2. Day 60 EQ VAS Results**

| | | N | Mean (SD) | Median (IQR) |
|---|---|---|---|---|
| **Antiviral domain** | | | | |
| | No antiviral therapy | 456 | 69.5(14.9) | 70(60-80) |
| | Paxlovid | 241 | 64.4(16) | 65(50-75) |
| | Azvudine | 157 | 67.3(15.4) | 70(60-80) |
| **Immune modulation domain** | | | | |
| | No immune modulator | 629 | 67.8(15.9) | 70(60-80) |
| | α-thymosin | 116 | 68.6(14.8) | 70(60-80) |
| | Baricitinib | 39 | 65.1(12.7) | 65(55-75) |
| | IL-6 receptor antagonist | 38 | 60(14.9) | 52.5(50-70) |
| **Immunoglobulin domain** | | | | |
| | No immunoglobulin | 788 | 67.3(15.8) | 70(60-80) |
| | Intravenous immunoglobulin | 70 | 65.4(14) | 70(50-75) |
| **Anticoagulation domain** | | | | |
| | Thromboprophylaxis | 528 | 67.4(15.1) | 70(55-80) |
| | Therapeutic anticoagulation | 121 | 62(16.3) | 60(50-70) |
| **Antiplatelet domain** | | | | |
| | No antiplatelet agent | 234 | 72.5(15.1) | 75(60-85) |
| | Antiplatelet agent | 228 | 65.3(13.8) | 65(55-75) |
| **Glucocorticoid domain** | | | | |
| | No glucocorticoid | 224 | 69.1(15.3) | 70(60-80) |
| | Glucocorticoid | 719 | 66.4(15.6) | 70(55-80) |

SD denotes standard deviation; IQR, interquartile range

**Table 3. Day 60 Disability Categories**

| | | Complete Disability n/N(%) | Severe Disability n/N (%) | Moderate Disability n/N (%) | Mild Disability n/N (%) | No Disability n/N (%) |
|---|---|---|---|---|---|---|
| **Antiviral domain** | | | | | | |
| | No antiviral therapy | 1/458(0.2) | 175/458 (38.2) | 112/458 (24.5) | 155/458 (33.8) | 15/458 (3.3) |
| | Paxlovid | 3/236(1.3) | 53/236 (22.5) | 56/236 (23.7) | 107/236 (45.3) | 17/236 (7.2) |
| | Azvudine | 1/157(0.6) | 39/157 (24.8) | 45/157 (28.7) | 66/157 (42) | 6/157 (3.8) |
| **Immune modulation domain** | | | | | | |
| | No immune modulator | 2/629(0.3) | 201/629 (32) | 144/629 (22.9) | 249/629 (39.6) | 33/629 (5.2) |
| | α-thymosin | 2/117(1.7) | 35/117 (29.9) | 38/117 (32.5) | 38/117 (32.5) | 4/117 (3.4) |
| | Baricitinib | 0/40(0) | 9/40 (22.5) | 5/40 (12.5) | 25/40 (62.5) | 1/40 (2.5) |
| | IL-6 receptor antagonist | 0/36(0) | 5/36(13.9) | 6/36 (16.7) | 23/36 (63.9) | 2/36 (5.6) |
| **Immunoglobulin domain** | | | | | | |
| | No immunoglobulin | 3/785(0.4) | 236/785 (30.1) | 190/785 (24.2) | 323/785 (41.1) | 33/785 (4.2) |
| | Intravenous immunoglobulin | 2/71(2.8) | 20/71(28.2) | 16/71(22.5) | 29/71(40.8) | 4/71(5.6) |
| **Anticoagulation domain** | | | | | | |
| | Thromboprophylaxis | 4/527(0.8) | 162/527(30.7) | 125/527(23.7) | 209/527(39.7) | 27/527(5.1) |
| | Therapeutic anticoagulation | 0/120(0) | 22/120(18.3) | 30/120(25) | 59/120(49.2) | 9/120(7.5) |
| **Antiplatelet domain** | | | | | | |
| | No antiplatelet agent | 2/235(0.9) | 128/235(54.5) | 53/235(22.6) | 46/235(19.6) | 6/235(2.6) |
| | Antiplatelet agent | 0/228(0) | 53/228(23.2) | 61/228(26.8) | 103/228(45.2) | 11/228(4.8) |
| **Glucocorticoid domain** | | | | | | |
| | No glucocorticoid | 0/223(0) | 74/223(33.2) | 57/223(25.6) | 81/223(36.3) | 11/223(4.9) |
| | Glucocorticoid | 5/717(0.7) | 205/717(28.6) | 175/717(24.4) | 298/717(41.6) | 34/717(4.7) |

#### Table 4: Day 60 Disability Comparison

| | | N (survivors at day 60) | WHODAS score, median (IQR) [N] | Adjusted odd ratio (95% CrI) | P value |
|---|---|---|---|---|---|
| **Antiviral domain** | | | | | |
| | No antiviral therapy | 458 | 86.5(59-132) | 1.00(reference) | |
| | Paxlovid | 236 | 109(73-147) | 0.45(0.32,0.62) | <0.001 |
| | Azvudine | 157 | 102(71-141) | 0.68(0.47,0.97) | 0.034 |
| **Immune modulation domain** | | | | | |
| | No immune modulator | 629 | 98(65-144) | 1.00(reference) | |
| | α-thymosin | 117 | 92(67-124) | 1.43(0.94,2.18) | 0.095 |
| | Baricitinib | 40 | 135.5(79-156) | 0.68(0.33,1.37) | 0.282 |
| | IL-6 receptor antagonist | 36 | 120(99-133) | 0.75(0.25,2.30) | 0.621 |
| **Immunoglobulin domain** | | | | | |
| | No immunoglobulin | 785 | 99(65-141) | 1.00(reference) | |
| | Intravenous immunoglobulin | 71 | 102(67-143) | 1.68(1.00,2.82) | 0.052 |
| **Anticoagulation domain** | | | | | |
| | Thromboprophylaxis | 527 | 99(65-141) | 1.00(reference) | |
| | Therapeutic anticoagulation | 120 | 114.5(81-156) | 0.65(0.44,0.96) | 0.03 |
| **Antiplatelet domain** | | | | | |
| | No antiplatelet agent | 235 | 69(56-102) | 1.00(reference) | |
| | Antiplatelet agent | 228 | 107.5(75-144.5) | 0.62(0.38,1.02) | 0.06 |
| **Glucocorticoid domain** | | | | | |
| | No glucocorticoid | 223 | 96(61-129) | 1.00(reference) | |
| | Glucocorticoid | 717 | 101(68-145) | 1.16(0.84,1.60) | 0.355 |

IQR denotes interquartile range; CrI, credible interval

Odds ratio parameters are for the analysis of WHODAS disability category. An OR<1 indicates reduced disability.

P value is computed from an ordinal mixture mode
